# A qualitative exploration to inform an oral health training for disability care workers in Burkina Faso

**DOI:** 10.1101/2023.08.02.23293545

**Authors:** Ave Põld, Dan Filwendé Kientega, Jocelyne Valérie Garé, Michael Lorenz

## Abstract

**Introduction:** Significant inequalities in oral health needs exist among people with intellectual and developmental disability in low-income settings such as Burkina Faso.

**Aim:** To explore enablers and barriers to the creation of an oral health training for caregivers of children with disabilities at specialized centres in Ouagadougou.

**Method:** This was a formative study informed by the Theoretical Domains Framework (TDF) using qualitative methods comprising observations at five centres for disability and semi-structured interviews with 14 caregivers.

**Results:** A successful training facilitation to caregivers must account for resources available in centres, part of the training must be dedicated to healthy diets and provide ideas for lowering sugar consumption and the training must empower caregivers with practical skills and tools to manage regular toothbrushing and identification of child oral health needs.

**Discussion:** These observations and interviews enabled us to gather valuable and unique perspectives into the everyday functioning of centres for disability, and the professional and personal role of caregivers working with children with disabilities in Ouagadougou.

**Implications for practice:** Teams planning oral health promotion activities in low resourced settings for vulnerable population groups can benefit from the methodology and results of this research for ensuring their interventions are appropriate and relevant.

**Relevance statement:** This is a unique field study conducted in a scarcely researched area of caregiving practices for children with disabilities in a low-income country, Burkina Faso. Results from the disability centre observations and interviews with local caregivers are of great value to any team planning health projects in similar low-resourced settings. Psychiatric and mental health nursing practices are highly context-dependent, thus using proposed qualitative methods can help to ensure that planned interventions are appropriate and relevant.

**Accessible summary:** What is known on the subject?

- People with disabilities often suffer from oral disease and lack opportunities to access oral care, especially in low-income countries such as Burkina Faso.
- Their caregivers and nurses lack training in oral health, necessary resources, and motivation to take care of oral hygiene and diet.

**What the paper adds to existing knowledge?:**

- This paper provides a detailed description of the everyday activities and functioning of disability care centres in Ouagadougou, Burkina Faso and insider perspectives related to the struggles, joys, and motivation of carers in performing their work activities.
- This paper adds new knowledge about the reality of disability care in Burkina Faso and gives practical information to teams planning health promotion activities in similar contexts, focusing primarily on oral health promotion and training.

**What are the implications for practice?:**

- Dedicating effort to improving the oral health of people with disabilities depends largely on nurses and carers having necessary resources including materials, practical skills, time and support to be empowered in their duties.
- Results from the disability centre observations and interviews with local caregivers are of great value to any team planning health projects in similar low-resourced settings with mental health nurses or carers.

## Background

Significant inequalities in health can be seen among people with intellectual and developmental disability ranging from a higher frequency of emergency hospital admissions (Hosking et al., 2017) to high rates of unmet oral health needs (Petrovic et al., 2016). People with disabilities often face a deteriorated oral health status and lack opportunities to access oral care (Wilson et al., 2019). In a low-resourced setting such as Burkina Faso where there is a high oral disease burden among people with disabilities, prevention and health promotion carry an important role in mitigating the need for curative oral health interventions (Lai et al., 2014).

Burkina Faso was one of the first West African countries to provide schooling for children and adolescents with disabilities starting from as early as the 1970s (Danielle Tan, 2020; Peyraud, 2018). Starting from specialised schools for children with specific types of disabilities, they gradually started integrating them to ordinary schools. Despite this history, educating children with disabilities still remains a challenge as most of the schools are expensive private institutions with high tuition fees that are located in the capital Ouagadougou, not in rural areas where 80% of people with disabilities live (Danielle Tan, 2020).

The first census of children with disabilities done in Burkina Faso in 2013 revealed that families are interested in providing their children high-quality education, yet managing their healthcare needs come first (Danielle Tan, 2020). Healthcare and well-being services for the disabled include a vast array of assistive products and inclusive supplies such as wheelchairs, spectacles, walking aids and modified educational materials (UNICEF, n.d.). Given the limited resources available at centres and schools for disabilities in Burkina Faso, procuring such products, training the staff, and ensuring children’s access to regular medical care including oral health care, remains challenging.

The importance of conducting formative research to guide implementation design and evaluation, is essential when conducting global health research (David Beran et al., 2018; Stetler et al., 2006). This study presents a qualitative evaluation focused on a scarcely researched area-improving the oral health of children with disabilities in a low-income country, Burkina Faso. The results of this study will be used to inform the creation and implementation of an oral health training for participating caregivers.

## Methods

### Theoretical framework

The qualitative data collection was informed by the Theoretical Domains Framework (TDF) (Cane et al., 2012). The TDF was selected as it suited best our aim of exploring behavioural determinants related to oral health related activities among participating caregivers.

### Study design

This is a formative study using qualitative methods comprising qualitative observations at five centres for disability and semi-structured interviews with 14 caregivers, in Ouagadougou. The current paper informs the development of an oral health training for caregivers.

### Study population and recruitment

Caregivers were presented an information sheet about the study and asked to sign a consent form. Full consent to participate in the study activities was gathered from all caregivers in the official language (French) with explanations provided in French and Mooré. They were approached by research team members and selected based on availability to participate in the interviews. Enrolment to the study was voluntary and participants could withdraw from the study at any given point.

### Data collection and tools

A field research team was assembled consisting of six local dental students. The students received regular trainings before each new phase of data collection by the lead researchers. The decision to train dental students to perform the data collection was guided by the need to have a field research team who is local, speaks French and the local language (*Mooré*) and can easily be accepted by the caregivers and children at the centres. By making the children and caregivers more used to having dental students regularly in the centres, we also aimed to minimise the Hawthorne effect during the data collection activities performed in centres (Goodwin et al., 2017).

#### a) Qualitative observations

An observational guide was established based on literature addressing the research objectives and relevant TDF domains before the start of the field activities. After visits to the disability centres and internal discussions within the research team, the observational guide was slightly modified to better capture areas related to the context in which the caregivers work at each centre.

Observations were conducted in 5 of a total of 6 participating centres. One of the centres was omitted from the observation as caregivers working there uniquely perform house visits to children with disabilities. Three pairs of research team members performed 3–4-hour observations at each centre.

#### b) Semi-structured interviews

Following the centre observations, the initial interview guide was modified to further investigate aspects captured during observations.

The team conducted 2-3 one-to-one interviews in each centre. They worked in pairs where one person interviewed and the other took notes and captured voice recordings using Kobo Toolbox (*Kobo Toolbox*, 2022). A total of 14 interviews were conducted with caregivers from 6 centres, each interview’s duration being 15-20 minutes.

### Analytical approach

The team members conducting observations wrote detailed observational reports based on the notes gathered using Kobo Toolbox from centres. Interviewers performed a verbatim transcription of the audio recording and cross-validated with the collected notes. The data was then translated from French to English for coding and thematic analysis using MAXQDA 2022 version 22.4 (VERBI Software, 2022). The first version of the coding scheme included mainly TDF domains (v2) and some inductive codes that emerged during the preliminary coding of 2 interviews. The codebook in MAXQDA included each code’s name, the decision rule for what to include or exclude, and an example.

2 interviews were randomly selected to create the reliability sample (≈ 14%) whereas the reliability sample was a subset of the full sample. The units of analysis were chosen to be paragraphs of the interviews as the interviews were short and all content of the interviews was intended to be coded. Both coders started with coding the same 2 interviews followed by assessing the intercoder agreement using MAXQDA and negotiating discrepancies and updating some code descriptions. In each discrepant case, we kept track of whether we achieved reconciliation and if so, which coder deferred to the other (Campbell et al., 2013).

Once a good agreement (more than 85% was reached), coders continued coding the remaining interviews followed by a negotiation and calculation of the intercoder agreement. During each round of coding, Kappa scores were also calculated for assessing intercoder reliability (Brennan & Prediger, 1981). By following this iterative process of unitizing, coding, discussing coding discrepancies, and refining codes and code definitions, intercoder agreement gradually improved leading to an overall agreement of 97,4% across all coded interviews.

The initial steps in constructing themes were aided by “Code Maps” and “Code Relations Browser”-MAXQDA visualization tools enabling to analyse which codes were assigned most, also the co-occurrence of and similarity between codes. Centre-level thematic analysis of observation and interview data was then performed where the most assigned and co-occurring codes were grouped together, the corresponding coded paragraphs analysed and further grouped into subthemes.

### Rigour and validity methods

Rigour and validity were addressed in different ways throughout the study design (Creswell & Miller, 2000). The interview sample was chosen from the target audience group by including 2-3 caregivers from each centre for disability. Multiple data sources were used to triangulate findings from qualitative observations and one-to-one interviews. To ensure a high validity of findings, the research team was rigorously trained in performing, recording, and reporting observations and interviews using standard procedures.

## Results

### Changes to the TDF and supplemental codes

Minimal changes were made to the definitions of two TDF domains. An explanation was added to the memo of the domain “Environmental context and resources” explaining that the code should only be used when the described context has a potential impact on the caregiver’s work. A clarification was also added to the memo of the domain “Knowledge” specifying that the code should also be used in cases of lack of knowledge or no knowledge about a topic covered during the interview such as toothbrushing. A set of supplemental inductive codes and their descriptions have been presented in Table 1.

**Table 1.** Supplemental interview codes with descriptions.

| **Code label** | **Code description** |
| --- | --- |
| toothbrushing_center | We asked the caregivers to give us suggestions on how it would be best to do the toothbrushing in their centre. We use this code when coding sections where they provide any inputs on how, when, where, etc. to do toothbrushing activities in the centre. In addition, information on what is the current or expected role of the caregiver after the training in relation to brushing. Includes any previous information about any toothbrushing related activities in the centre either before or now. |
| disability | This code is used in sections where caregivers talk about the disabilities of the children.<br>This may include referring to difficulty in their work due to children being disabled, or learning/teaching difficulties due to the children being disabled, also when they specify what types of disabilities the children have in their centre, etc. |
| center_org_meal | To describe when and how meals are organized by the centres- time, place, etc. Whether there are issues that children have with eating (in this case if they mention also disability, then it should be coded with both the codes "center__org_meal" and "disability"). |
| activities | This code should be used if caregivers talk about any activity they perform with the children. This may include motoric training, psychomotor training, writing, activities where they make pearl necklaces, play with the balloon, sing, do speech therapy, communication, also teaching them practical skills such as washing hands. Also, decisions related to how and what regular activities with children are done can be coded using "activities". |
| center_providedmeal | This code is to be used when caregivers describe how meals are provided in their centre: what foods and drinks. This code is NOT to be used if the children eat their own meals. If the answer is about what food the children eat (own or centre food). IF it is not clear who provides the meal or drink, we use both "center_providedmeal" and "own_meal". |
| own_meal | This code is to be used in cases where centres do not offer food to the children, but the children eat meals their parents packed for them. Also, for coding the answer where caregivers specify that the parents DO NOT give additional food for the kids. IF it is not clear who provides the meal or drink, we use both "center_providedmeal" and "own_meal". |
| nobrushing | This code is used in response to the question asking about toothbrushing activities in the centre. If there is no toothbrushing being organised by the centre, then this code should be applied. |

### Major themes for oral health promotion activities

#### 1. The setting: the six centres

Two of the centres included in this study are non-governmental organizations (NGOs) and four are associations. All participating centres differ in how they are managed with four centres acting as day-care centres for disabled youth, one performing house visits and one being both a residential home and a day-care centre. There are on average 50-100 children and adolescents per centre. There are different levels of human resources involved in each centre including administrative, caregiving, security and catering staff ranging from a total of 5 to 25 people working per centre.

Most of the disability centres include youth with different types of disability, and often they are mixed in classes. That means that caregivers must manage children with autism spectrum disorders, various syndromes such as Down’s syndrome, encephalopathy, and other disabilities in the same classroom. The children are allocated based on their capabilities, not their disability, to classes by nurses, doctors, and psychologists. In most cases while working with children, caregivers find out more about them and can reassign pupils where necessary.

#### 2. Providing care and education for children with disabilities

##### Daily activities of caregivers

Caregivers facilitate different activities with the children throughout the day. Depending on the centre, they target enhancing various cognitive and motoric skills of children or do simply pastime activities. In one institution there are preparation classes where children are taught to write letters and numbers, they also work on communication and motoric skills depending on the needs of the child. In addition to managing with multilingual verbal communication in Mooré, Dioula and French, caregivers also engage on a regular basis in non-verbal communication with children who can’t speak. As one interviewed caregiver described the situation, “*There are some that cannot speak and others who can speak but do not know how to speak*.”

In one centre the day begins with a session of “free play” where children play with various toys, this is followed by beading to train their fine motoric skills, then a game to learn names by throwing a ball between each other and finishing with listening to music and singing along. In another centre children are divided based on their age into 2 groups with similar activities including beading and listening to music. One of the main differences noticed during observations is the difference in resources and the activity of carers. In the first centre they intervene only if there are arguments between children but in the second institution they are actively engaged and coordinate continuously the activities and provide each other support.

One of the six centres is unique as it also hosts children with disabilities on a permanent basis. This centre also manages its activities differently in a similar manner to family life where children prepare food together, help with gardening work, have workshops and other everyday activities.

> An interviewee description about the centre, “*We welcome children who, according to the law, would not be able to survive if we did not take them. Some don’t have parents. Others have families but it doesn’t work in those families. Some of them, if they were not welcomed here, it is not certain that they would survive*.”

There is also one centre where caregivers work by performing family visits, examinations in hospitals or rehabilitation centres for the disabled. They are working closely with most of the public and private health structures in the capital thus also facilitating consultations and screenings at schools.

> A caregiver on the importance of sensitization activities, “*It’s a moment of sharing with parents and teachers. They discover what disability is. Especially teachers who can punish children thinking they are stupid when they have a disability. [It is important to] Do early screening*.”

##### The struggles of everyday work

During the interviews we also tried to investigate some difficulties that caregivers face in their everyday work. Several of them mentioned that some children are often difficult to manage, and they tend not to obey to orders and not engage in activities planned for them. In most cases the caregivers also felt frustrated because of a lack in staff to manage the children with special needs. There are occasional situations where some children have seizures or act impulsively due to their disability and with big groups of children it is often extremely difficult to always take care of the well-being of everyone.

> Another challenge described by interviewed caregivers are unengaged parents, “*For them [parents], it’s like a garage-they come and do everything to get rid of the child, whereas there are instructions that you can give to the parents if they agree to do it, it can help the child*.”

##### The joys of being a caregiver

> “*The discipline of the disabled children in this centre is remarkable, the children are in perfect cohesion, and help each other with activities. There is a notion of living together among people with intellectual disabilities.”* (An observation)

Our team observed that in all centres the caregivers are dedicated in their work despite the lack of resources and vast struggles that they often face. All the 14 interviewed caregivers enjoyed their work at the centres mainly because of the children they take care of. They emphasised the human aspect of their work in trying to improve the lives of the children with disabilities and give them some enjoyment in life. One of the interviewees had been working since 2003 in the centre and explained that without loving the kids it would be impossible to do this job for such a long time. Another theme that echoed through was the importance of patience and time when working with these children as they may provoke and need more effort to achieve expected learning outcomes. After working longer in the centre, the carers become friends with the children and as the connections are tight, they often sense having two families-one at home and one at work. The process of learning to communicate with the children takes time but once achieved, gives back on both emotional and professional levels, “*Communicating with them is like learning at school: as you go along you learn things that you might like*.”

#### 3. Planning for action to improve oral health

##### Enablers and restrains for implementing toothbrushing

We gathered information about the overall feasibility to plan toothbrushing activities in each centre and investigated what resources and tools the carers need for a successful implementation. Caregivers were asked to advise us on when and where it would be best to conduct toothbrushing. In some cases, caregivers said that the centre bathrooms are not suitable for toothbrushing but offered to bring a big water container with a tap to manage. Each caregiver gave suggestions on the ideal time of the day to do toothbrushing which mostly was either before or after a meal.

None of the carers facilitated toothbrushing activities at the start of our project, with the only exception being the centre where children also live. When asked about toothbrushing from interviewees, the responses ranged from teaching children about personal hygiene such as washing hands and eyes to instructing them to brush their teeth at home before coming to school. Some shared stories from their experiences with brushing the children’s teeth where *“there are some kids who grab the brush thinking that it is food, there are others who have broken the stem of the brush and do not allow anyone near their mouths.”* If children display a bad behaviour with certain supervisors, then they try to change the carer to best fit the mood of the child.

When discussing previous oral health activities, an interviewee mentioned “the big Olympic association that often organizes oral health activities and that we participate in”, the Burkina Faso country office for the Special Olympics (Special Olympics, 2023). As most oral health promotion is highly dependent on external partners, it poses huge challenges in regularly renewing brushing material for the children as they “sometimes get brushing kits here, but not all the time”. Some carers also highlighted the need to get the children’s teeth treated as some of them already have cavities.

> *“We who live with these children see that there are some children who need a consultation to treat cavities. With them, when we use brushes, they deteriorate quickly, so if we can have pastes and brushes for them, it will help us a lot.”*

##### The impact of diet on oral health

Another focus area for planning the training was nutrition and diet as unhealthy eating behaviours impact the incidence of dental caries and gingivitis. During observations and interviews we learned about the frequency and type of meals provided to the children.

The frequency of meals in most centres was restricted to 2-3 times a day with 2-4 hour breaks in between. Several caregivers mentioned that for the morning snack parents either pack something for their children or the centre offers provides this snack. In most cases these are sugary foods such as cakes and cookies that are taken with sweet juice (bissap), chocolate milk, or milk. Usually water is available, but children tend to bring their favourite drinks with them which often include various juices. There is usually no snacking in between meals but in one centre food is used as reinforcers to motivate children.

> A caregiver explaining the usage of “reinforcers”, *“Here sometimes with the children, we need food that we call “reinforcers” for example if the child does not adhere to the activity. It can be a food or a juice, we try to bring him to discipline by giving him just a small amount of his reinforcers. This principle is regulated by the staff and not the child.”*

The lunches prepared by the centres are warm local foods such as rice with sauce or beans. Where necessary, caregivers assist children in eating but most of them manage themselves according to the interviewees. Only one caregiver mentioned that they try to be careful in not allowing children to eat too much as it is bad for their wellbeing, but most centres give food to the children until they say or show that they are full.

##### Expectations towards the oral health training

Most caregivers expressed wanting to learn about oral hygiene to prevent teeth from becoming ill. Several of them mentioned that the training would help them both professionally and personally to take better care of their own oral health and that of their family. The motivation of caregivers and hesitancy to pick up additional oral health commitments also echoed in an interview, *“If I attend the training, it is to discover. Otherwise, I am not an oral specialist. I have already had toothaches, I went to see a doctor, I was treated.”*

All the caregivers knew what resources they are lacking to start regular toothbrushing: training, toothpaste, toothbrushes and in some cases better access to water. Assistance and advice on techniques to facilitate toothbrushing of children were expected to be covered in the training as well. As one carer explained, “*The brushes must be adapted, that there is a way of brushing but all that I do not know. I want to learn all this.”*

It was very reassuring hearing from multiple caregivers that once they receive the oral health training and basic resources for toothbrushing, they will easily start implementing it as part of their work, *“If we have the necessities we can motivate them, if we have the products, we can mentor them.”* Such positive feedback from caregivers is expected to support the upcoming training and implementation activities and will be evaluated in the upcoming phase of research.

## Discussion

Based on workplace observations and one-to-one interviews we identified three major themes to be considered for a successful training facilitation to caregivers. These included accounting for available resources in centres, dedicating time to cover healthy diets, providing practical tips for lowering sugar consumption and empowering caregivers with practical skills and tools to manage regular toothbrushing at centres.

The training must account for the different resources available in centres to increase caregiver motivation to apply learnings to their everyday work with children with disabilities. People with disabilities struggle in accessing health promotion interventions aimed to improve the self-management of their health and wellbeing (Jobling & Cuskelly, 2006; Rimmer et al., 2014). They are highly dependent on their caregivers for support and guidance, yet often carers are faced with multiple challenges in their jobs thus also impacting the outcomes for children. As the classes are mixed and comprise children with various types of disability, their needs are often very specific, and caregivers may lack human resources, skills or knowledge to tackle all situations. The reported lack of personnel could limit the successful implementation of oral health activities to everyday practise despite all interviewed caregivers having stated that once they receive training and toothbrushing material, they would proceed to practise oral health activities at work.

Previous studies have emphasised the cost of caregiving as a challenge in providing care (Ndadzungira, Allan., 2016). We also witnessed during centre observations how more well-off centres had smaller groups of children, more caregivers, more assistive technology thanks to more funding from parents and sponsors. As all the centres are funded to some extent by external sponsors, various dependencies exist when procuring materials and equipment and planning activities with the children. As one of the centre managers mentioned, due to the COVID-19 pandemic and the surge of terrorism in the country, multiple sponsors find it difficult to provide support and to travel to Burkina Faso, thus having a huge negative impact on the overall care provision for children and adolescents with disabilities. Caregivers and centre coordinators stated that they work with mainly foreign donors and mobilizing national donors is complicated for them. There is a need for more technical and financial partnerships yet establishing and maintaining them remains challenging.

A part of the training must be dedicated to healthy diets and provide ideas for lowering the sugar consumption of children in each centre. We learned that specific times for meals are respected in all centres but in most cases, parents pack sugary snacks and sweet drinks for their children and centres also offer juices and chocolate milk as drinks instead of just plain water. This might be due to a lack of safe drinking water in the centres or a relatively high price of bottled water forcing children to drink sweet drinks (Atin Prabandari, 2019; Mosites et al., 2020). Another interesting behaviour that some caregivers mentioned, was the use of food to reinforce good behaviour among children. It is common for children with autism spectrum disorders to exhibit unusual eating patterns such as selective eating (Bandini et al., 2010). They tend to prefer energy-dense foods such as sugary snacks, sugar-sweetened beverages and often refuse vegetables (Bandini et al., 2010; Evans et al., 2012). The use of snacks as incentives results in a high frequency of food intake, which can be harmful to the oral health of those children in care.

The training must empower caregivers with practical skills and tools to manage regular toothbrushing and better identify the oral health needs of children with disabilities. People with disabilities have higher rates of dental caries, more extensive periodontal destruction (Sasaki et al., 2004), significantly more missing teeth, and less restored teeth compared to the general population (Kebede et al., 2012; Lee et al., 2019). Their caregivers are usually not engaged in routine activities to prevent oral disease and they often underestimate the oral pain of people with disabilities thus delaying their access to dental care (Lee et al., 2019). We also found that in the centres that children attended during the day, there was no toothbrushing and caregivers were often even surprised at the question, not knowing how to respond. At the residential centre where children brushed their teeth, caregivers were also more aware of their oral health situation stating that several children need dental care as they see while assisting them in toothbrushing, that they have cavities. This finding supports the need to train caregivers to identify oral disease and to support the provision of good oral hygiene for all children. In Burkina Faso where based on WHO 2018 data there are just 91 dentists and 185 dental therapists for a population of 22,1 million, there is a severe lack of oral care providers with a vast majority being private and too expensive for most of the population to even access (WHO, 2022; World Bank, 2022). The Burkinabe government implemented in 2010 a law on the protection and promotion of the rights of persons with disabilities stating that any person with a disability benefits from free consultations, care, complementary examinations, medicines and hospitalization in public health structures (The Parliament of Burkina Faso, 2010). The extent to which this law is being implemented is difficult to assess but based on the 6 centres we conducted our research at, they all receive external non-national funding, the orthopaedic and therapeutic devices are old, mainly donated from Europe and need renewing, the children are not regularly taken to public health facilities to the dentists or dental nurses although there is a perceived need noted by the interviewed caregivers.

The main limitations of the study were related to observation reporting, interview sampling, respondent bias and TDF use. As observations and interviews were conducted by different pairs of dental students, the reporting quality slightly varied. The participants for interviews were self-selected by the research team members based on availability. This could have led to a biased selection of caregivers as demographic characteristics were not considered when enrolling participants. The interviews were conducted in French but in some cases, there were communication problems when conducting the interviews as some caregivers were native speakers of Mooré. During the analysis phase the TDF domains were used as the starting point in coding the observations and interviews, yet minor modifications were made to some domain descriptions and inductive codes targeting oral health specific topics were added thus decreasing the weight of the TDF in the overall analysis process.

## Conclusion

This is the first research study targeting oral health improvement for people with disabilities that has been conducted in Burkina Faso. We identified strong interest among caregivers to attend an oral health training and receive materials for implementing toothbrushing activities in each centre. These observations and interviews enabled us to gather valuable and unique perspectives into the everyday functioning of these centres for disability and the professional and personal role of caregivers working with children with disabilities in Ouagadougou.

## Data Availability

All data produced in the present study are available upon request to the authors.

## Acknowledgements

We sincerely thank our local data collection team for their hard work and dedication (Dora Marie Paule Gueye, Fayshal Abdoul Kiendrebeogo, Hanatou Zampou, Laure Denise Gampine, Léonard Souka Kabore, Mohamed Fahad Issoufa Madi). We are also grateful for the support of all participating disability centre coordinators and caregivers and of the Dental Department of the Joseph-Ki Zerbo University.

